# Maternal Urinary Phthalate Metabolites are Associated with Lipidomic Signatures Among Pregnant Women in Puerto Rico

**DOI:** 10.1101/2021.07.19.21260700

**Authors:** Pahriya Ashrap, Max T. Aung, Deborah J. Watkins, Bhramar Mukherjee, Zaira Rosario-Pabón, Carmen M. Vélez-Vega, Akram Alshawabkeh, José F. Cordero, John D. Meeker

## Abstract

**Background:** Phthalates have been reported to alter circulating lipid concentrations in animals, and investigation of these associations in humans will provide greater understanding of potential mechanisms for health outcomes.

**Objective:** to explore associations between phthalate metabolite biomarkers and lipidomic profiles among pregnant women (n = 99) in the Puerto Rico PROTECT cohort.

**Methods:** We measured 19 urinary phthalate metabolites during 24-28 weeks of pregnancy. Lipidomic profiles were identified from plasma samples by liquid chromatography-mass spectrometry-based shotgun lipidomics. Relationships between phthalates and lipid profiles were estimated using compound-by-compound comparisons in multiple linear regression and dimension reduction techniques. We derived sums for each lipid class and sub-class (saturated, monounsaturated, polyunsaturated) which were then regressed on phthalates. Associations were adjusted for false discovery.

**Results:** After controlling for multiple comparisons, 33 phthalate-lipid associations were identified (q-value<0.05), and diacylglycerol 40:7 and plasmenyl-phosphatidylcholine 35:1 were the most strongly associated with multiple phthalate metabolites. Metabolites of di-2-ethylhexyl phthalate, bis(2-ethylhexyl) phthalate, dibutyl phthalates, and diisobutyl phthalate were associated with increased ceramides, lysophosphatidylcholines, lysophosphatidylethanolamines, and triacylglycerols, particularly those containing saturated and monounsaturated fatty acid chains.

**Significance:** Characterization of associations between lipidomic markers and phthalates during pregnancy will yield mechanistic insight for maternal and child health outcomes.

**Impact Statement:** This study leverages emerging technology to evaluate lipidome-wide signatures of phthalate exposure during pregnancy. Circulating lipids are critical for biological processes including inflammation, cell-to-cell communication, and metabolism. Therefore, lipid signatures of phthalate exposure provide insight into potential toxicological mechanisms. Characterization of these mechanisms are relevant for informing the etiology of maternal and children’s health outcomes.

## INTRODUCTION

Phthalates are a group of chemicals that are found in numerous commercially available products like food packaging, vinyl flooring, pharmaceuticals, and personal care products [1, 2]. According to biomonitoring reports, human exposure to phthalates is ubiquitous for children and adults [3, 4], including pregnant women and their fetuses due to transplacental passage of phthalates [5]. Prenatal and early-life exposure to phthalates has been related to adverse pregnancy events [6-11] and childhood developmental and cardiometabolic outcomes [12-16].

One of the proposed mechanisms of action and pathways involved in early life phthalate-induced adverse effects is disruption of lipids. Animal studies suggest that phthalates influence circulating lipid levels and lipid signaling [17-20], by interfering with peroxisome-proliferator active receptor gamma (PPARγ) [21, 22]. This points to the possibility that a similar mechanism is at play for humans. However, the relationship between prenatal phthalate exposure and lipid metabolism has rarely been investigated among humans. In the limited existing research on humans, the focus has been on the relationship between maternal phthalates and specific lipids such as cholesterol and triglycerides [23-25]. Application of lipidomic profiling (lipidomics) of global cellular lipids in a complex biological sample [26], a branch of metabolomics, may inform research on the discovery of new potential biomarkers of effect or patterns of change related to environmental exposures, including phthalates. Only one study to date has examined the association between maternal metabolomics, including lipid metabolites, and environmental phthalates in pregnant women (n = 115) from the Center for the Health Assessment of Mothers and Children of Salinas (CHAMACOS) cohort [27]. The authors found significant relationships between maternal phthalate exposure and plasma free fatty acids, lysolipids, ceramides, and triacylglycerols.

Our previous research in the Puerto Rico PROTECT birth cohort found associations between prenatal phthalate urine metabolite levels and increased odds of preterm birth [28, 29]. In the PROTECT cohort, we also assessed the relationship between maternal lipidomic profiles and adverse birth outcomes and observed numerous significant lipid signatures for spontaneous preterm birth [30]. In the current study, we sought to explore the association between maternal phthalate metabolite biomarkers and lipidomic profiles among pregnant women (n = 99) in the PROTECT birth cohort. In doing so, we contribute to the characterization of the potential mechanism of action for phthalate exposure. We also highlight the utility of lipidomics as a tool in environmental epidemiology in detecting intermediate endogenous biomarkers of phthalate exposures.

## METHODS

### Study sample

This study sample is an exploratory subset of the Puerto Rico PROTECT cohort. The PROTECT cohort began recruitment in 2010 through funding from the National Institute of Environmental Health Sciences Superfund Research Program. Study participants were recruited in the first or second trimester of pregnancy (median 14 weeks gestation). The research protocol was approved by the Ethics and Research Committees of the University of Puerto Rico and participating clinics, the University of Michigan, Northeastern University, and the University of Georgia. Inclusion criteria for recruitment included: participant age between 18 and 40 years; residence in the Northern Karst aquifer region; disuse of oral contraceptives three months before pregnancy; disuse of in vitro fertilization; and no indication of medical records for major obstetrical complications, including pre-existing diabetes. Prenatal spot urine and blood samples were collected during the third study visit at approximately 26 weeks of gestation. From the larger cohort of 2070 pregnant women, we randomly sampled 100 women with the goal of achieving a 1:2 case-control ratio of preterm birth, which included 31 cases and 69 controls. We created and applied inverse probability weights to account for the sampling approach [31].

### Urinary phthalate measurements

Urine was collected in polypropylene containers, divided into aliquots, and frozen at -80 °C until shipment overnight to the Division of Laboratory Sciences at the Centers for Disease Control and Prevention (CDC) for analysis. Urinary concentrations of 15 phthalates, two DEHTP, and two DINCH metabolites were measured using on-line solid phase extraction coupled with isotope dilution-high performance liquid chromatography-electrospray ionization-tandem mass spectrometry as previously described [32-34]. Measured phthalate metabolites comprised mono(2-ethylhexyl) phthalate (MEHP), mono(2-ethyl-5-hydroxyhexyl) phthalate (MEHHP), mono(2-ethyl-5-oxohexyl) phthalate (MEOHP), mono(2-ethyl-5-carboxypentyl) phthalate (MECPP), mono-benzyl phthalate (MBzP), monocarboxyoctyl phthalate (MCOP), mono-isononyl phthalate (MNP), mono-oxoisononyl phthalate (MONP), monocarboxynonyl phthalate (MCNP), mono (3-carboxypropyl) phthalate (MCPP), mono-ethyl phthalate (MEP), mono-n-butyl phthalate (MBP), mono-3-hydroxybutyl phthalate (MHBP), mono-isobutyl phthalate (MiBP) and mono-2-hydroxy-iso-butyl phthalate (MHiBP). Measured DEHTP metabolites comprised mono-2-ethyl-5-carboxypentyl terephthalate (MECPTP) and mono-2-ethyl-5-hydroxyhexyl terephthalate (MEHHTP) [35, 36]. Measured DINCH metabolites were cyclohexane-1,2-dicarboxylic acid monohydroxy isononyl ester (MHiNCH) and cyclohexane-1,2-dicarboxylic acid monocarboxy isooctyl ester (MCOCH) [33]. The sum of di-2-ethylhexyl phthalate metabolites (ΣDEHP) was calculated by adding the molar fractions of MEHP, MEHHP, MEOHP, and MECPP. To achieve unit comparability, ΣDEHP (nmol/ml) was multiplied by the molecular weight (MW) of MEHP (278.348 g/mol). The resulting units were ng/ml. Specific gravity (SG) was measured using a handheld digital refractometer (Atago Co., Ltd., Tokyo, Japan) at the University of Puerto Rico Medical Sciences Campus at the time of sample collection. Values below the limit of detection (LOD) were imputed with the metabolite-specific LOD/√2 [37].

### Lipidome measurement

Plasma lipids were extracted from blood plasma samples provided by women at the third study visit using a modified Bligh-Dyer method [38] using liquid-liquid extraction at room temperature after spiking with internal standards. Analysis of lipids was performed on reversed phase high-performance liquid chromatography (HPLC), followed by mass spectrometry (MS) analysis that alternates between MS and data dependent MS2 scans using dynamic exclusion in both positive and negative polarity and yields excellent separation of all classes of lipids. The lipids are quantified using Multiquant and normalized by internal standards. Measurements are semi-quantitative and are based on the relative abundance of peak intensities. Quality controls (QC) are prepared by pooling equal volumes of each sample, in addition to a well characterized plasma pools, and are injected at the beginning and end of each analysis and after every 10 sample injections, to provide a measurement of the system’s stability and performance as well as reproducibility of the sample preparation method [39].

Lipids were identified using the LipidBlast [40] library (computer-generated tandem mass spectral library of 212,516 spectra covering 119,200 compounds from 26 lipid compound classes), by matching the product ions MS/MS data. The method allowed us to measure 587 lipids belonging to 19 different lipid classes which include acylcarnitine (AcyICN), ceramides (CER), cholesterol esters (CE), diacylglycerols (DG), glucosylceramides(GlcCer), free fatty acids (FFA), fatty acid esters of hydroxy fatty acids (FAHFA), lysophosphatidylcholine (LysoPC), lysophosphatidylethanolamine (LysoPE), phosphatidic acid (PA), phosphatidylcholine (PC), phosphatidylethanolamine (PE), phosphatidylglycerol (PG), plasmenyl-phosphatidylcholine (PLPC), plasmenyl-phosphatidylethanolamine (PLPE), phosphatidylinositol (PI), phosphatidylserine (PS), sphingomyelin (SM), and triacylglycerol (TG). All individual lipids will be mentioned with the nomenclature as X:Y, where X is the length of the carbon chain and Y, the number of double bonds.

### Statistical Analysis

Inverse probability weighting of over-representation of preterm birth cases was applied to all statistical analyses presented in this analysis in order for our study to resemble the proportions of preterm birth in a general population [31]. 11 phthalate metabolites (MEHP, MEHHP, MEOHP, MECPP, MBzP, MCOP, MCNP, MCPP, MEP, MBP, MiBP) and the sum of DEHP metabolites (ΣDEHP) were included in the statistical analysis. Four phthalate metabolites (MNP, MONP, MHBP, and MHiBP), two DEHTP metabolites (MECPTP, MEHHTP), two DINCH metabolites (MHiNCH, MCOCH) had >15% data missing, therefore were excluded from the analysis.

The group sum of the individual lipids’ relative abundance that belong to each lipid class (n=19) were calculated to evaluate lipid classes. Considering lipid classes containing saturated, mono-unsaturated, and polyunsaturated fatty acids (PUFAs) may present different biological patterns, 45 lipid subgroups (saturated, mono-unsaturated, poly-unsaturated) were created by grouping lipids based on a priori knowledge of lipid class and the number of double bonds in each lipid species. Sum of lipids’ relative abundance were also created for the 45 sub-groups. Descriptive statistics for lipids were computed to examine their distributions. Spearman correlation coefficients were calculated between lipid class, lipid sub-groups and phthalates to visualize crude associations.

Multiple statistical strategies were used to assess relationships between lipid profiles and phthalates. The relative abundance of individual lipids, group sums, and sub-groups sums were log-transformed due to their skewed distribution. Subsequently, the log-transformed individual lipids, group sums, and sub-group sums were normalized among individuals (rows) by standard normal variate (SNV) normalization. Covariates were selected based on *a priori* considerations from literature review of factors that would influence both lipid concentrations and phthalate levels. Multiple linear regression models were fitted to evaluate the association between phthalates and the lipidome, adjusting for maternal age, maternal education, fetal sex, pre-pregnancy BMI, and weight gain during pregnancy. 1) Single pollutant models were performed for each log-transformed and standardized relative abundance of lipid (n=587) regressing on phthalate metabolites (n=12). False discovery rate (FDR) adjusted p-values (q-values), a commonly used method of adjusting for multiple comparisons in lipidomics studies, were used to account for multiple comparisons. 2) Sums for each lipid class (p=19) and lipid sub-group (p=45) (saturated, monounsaturated, polyunsaturated were then regressed on each phthalate metabolite (n=12). The results are presented as estimated percentage changes of lipids, lipid group sums, or sub-group sums with the corresponding 95% confidence intervals and p values per interquartile range (IQR) change in phthalate metabolite concentration.

## RESULTS

### Descriptive statistics

Demographic and health characteristics of the women in our study are reported in Supplementary Table 1. The mean age of study participants at the time of enrollment was 26.5 years (standard deviation = 5.7). A majority of participants (77%) had some level of higher education experience. In this sample, 44% of participants reported that they were unemployed, and 75% of participant household incomes were below $50k. Approximately 37% of participants had pre-pregnancy body mass index of greater than 25 kg/m^2^. Most participants did not report smoking (98%) or drinking (92%) during pregnancy.

### Phthalates and Lipidome-wide metabolite associations

**Figure 1** is a Manhattan plot indicating -log10(p-value) for phthalates in association with individual lipids. The dashed line indicates p value = 0.001. The 33 most significant associations after adjusting for false discovery rate (p value<0.001, q value<0.05) were annotated with the individual lipid in the figure. The Manhattan plot depicts that the lipid metabolites from the DG family were mainly associated with DEHP metabolites, while all of the significant signaling lipids from the CER class were found to be associated with MCNP, MCPP, MBP, and MiBP. Among these top associations, the most prevalent and significant signals were for lipid metabolite DG 40:7 in association with all four DEHP metabolites (MEHP, MEHHP, MEOHP, and MECPP), MCPP, and MBP. This was also confirmed on a feature level with a Euler diagram (**Supplementary Figure 1**) showing the overlap between significant lipid metabolite signals found across different phthalate parent compounds. Among all the lipids with significant signals, DG 40:7 overlapped within most of the ellipses. Another such signal is PLPC 35:1, where MEHHP, MEOHP, MECPP, MBP, and MiBP were significantly associated with this lipid metabolite from the PLPC family. In addition, urinary MECPP and MCPP concentrations were significantly associated with lipid metabolite LysoPE 18:0 from the LysoPE class. MBzP, MCOP, and MEP were not associated with any lipid metabolite in these individual models (**Figure 1 & Supplementary Table 2**).

**Figure 1.**
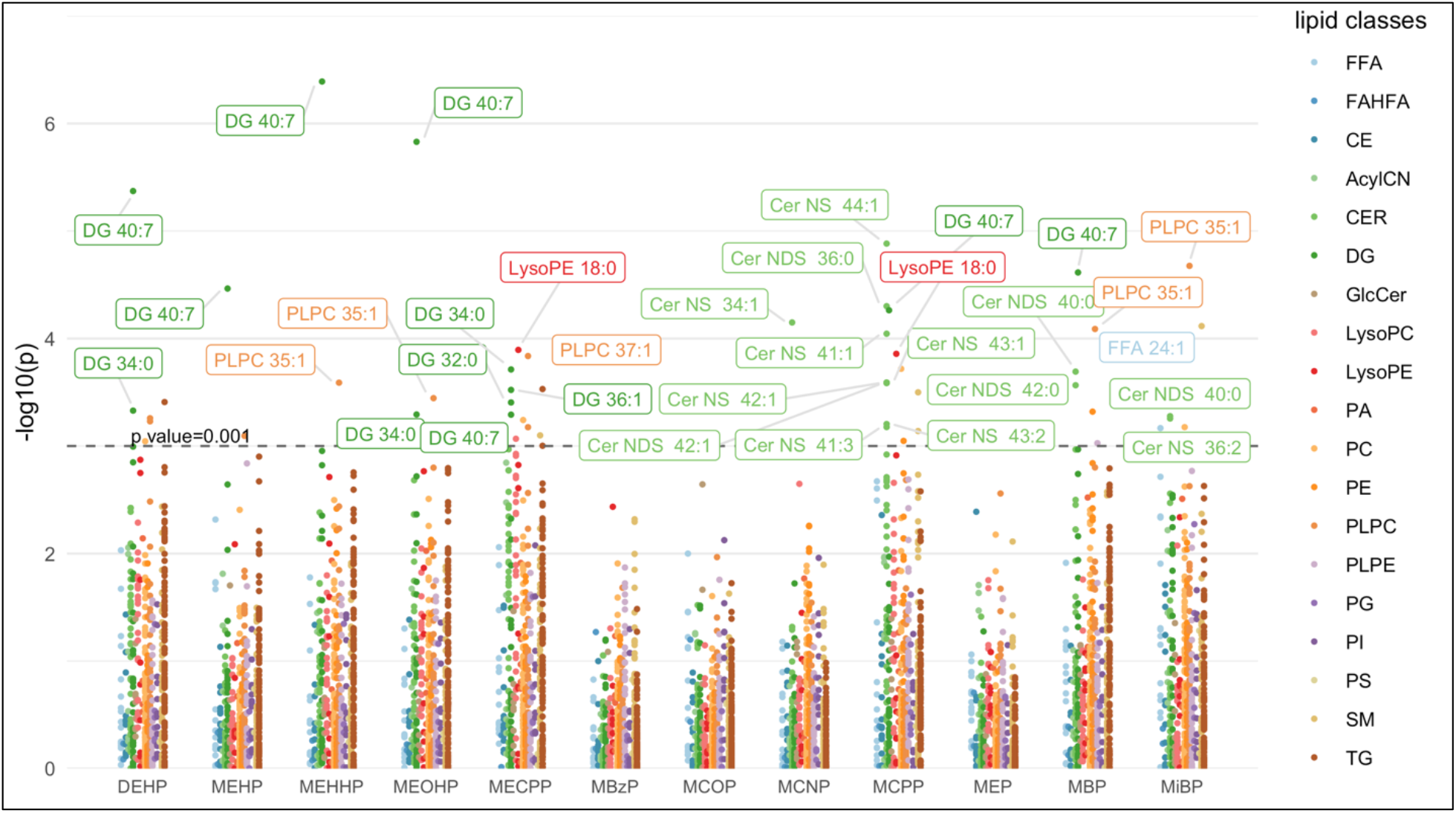
Manhattan plot showing individual lipids associated with urinary phthalate metabolite concentrations. Models were adjusted for maternal age, maternal education, fetal sex, pre-pregnancy BMI, and weight gain during pregnancy.

### Saturation subgroup associations with pregnancy phenotypes

A list of the individual lipids within each saturation group is reported in **Supplementary Table 3**. Associations between phthalates and the summation of individual lipid metabolites based on the degree of saturation in fatty acid hydrocarbon tails are presented in **Figure 2**. Significant associations after false discovery rate correction (p value<0.001 & q value<0.05) were annotated for lipid subclass with color indicating association direction, with red representing a positive association and blue indicating a negative association. The effect estimates and 95% confidence intervals of the significant associations are also presented in **Supplementary Table 4**. Nearly all significant associations between phthalates and lipid subgroups were positive. The three oxidative DEHP metabolites, MEHHP, MEOHP, MECPP, as well as the summary measurement ΣDEHP were all positively associated with increased saturated CER, saturated and mono-unsaturated LysoPE and PA, and saturated TG. Higher MECPP was additionally associated with mono-unsaturated CER as well as saturated and mono-unsaturated LysoPC.

**Figure 2.**
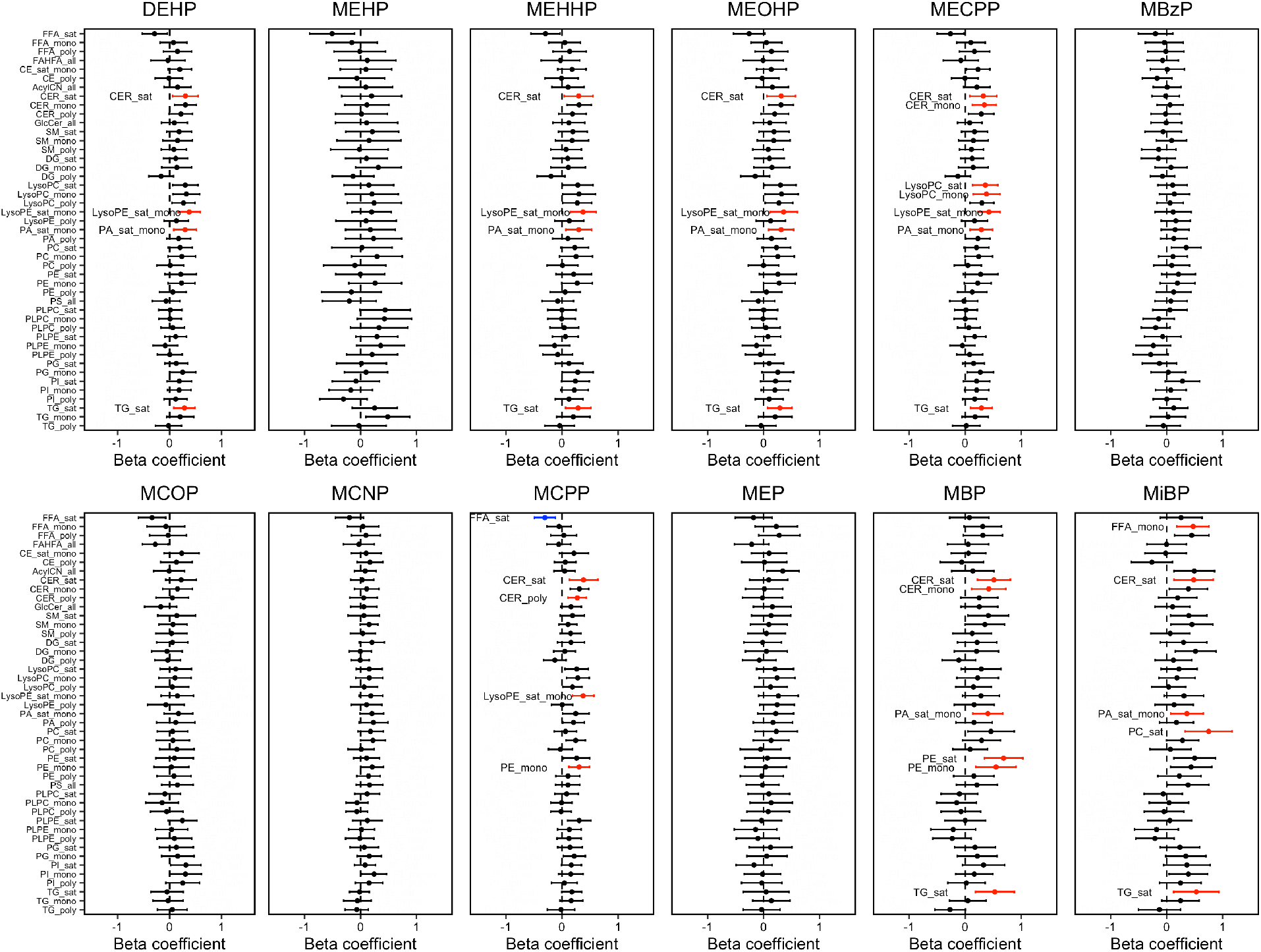
Percent change in lipid subgroup sum z-score associated with urinary phthalate metabolite concentrations. Effect estimates are presented as percent change (%) for IQR increase in exposure biomarker concentration^ab^. Models were adjusted for maternal age, maternal education, fetal sex, pre-pregnancy BMI, and weight gain during pregnancy.

For increased MCPP concentrations, we observed higher saturated and poly-unsaturated CER, saturated and mono-unsaturated LysoPE, and mono-unsaturated PE levels. The only negative association was found between increased MCPP concentrations and lower saturated FFA. The DBP metabolite MBP was associated with higher levels of saturated CER, PE, and TG, as well as mono-unsaturated CER, PA, and PE. The DiBP metabolite MiBP was associated with higher mono-unsaturated FFA and PA, in addition to saturated CER, PC, and TG. MBzP, MCOP, MCNP, and MEP were not significantly associated with any subgroup lipid class.

### Whole lipid classes associations with pregnancy phenotypes

The heat map in **Figure 3** shows the relationships between phthalate metabolites and lipid classes. Spearman ρ ranged from -0.22 to 0.27. Correlations between MECPP with LysoPC (ρ=0.27) and LysoPE (ρ=0.25) were among the most significant and positive, while the correlation between MEHP and FFA (ρ=-0.22) was the most negative. **Figure 4** shows forest plots of the associations between phthalates and whole lipid classes. Among the lipid classes including CER, LysoPC, LysoPE, and TG, we observed the following positive associations (p value<0.001 & q value<0.05) with ΣDEHP (LysoPE), MECPP (CER, LysoPC, LysoPE), MCPP (CER, LysoPE), and MiBP (TG) when clustering all lipid metabolites into their highest level of organization. **Supplementary Table 5** lists the effect estimates and 95% confidence intervals of these associations.

**Figure 3.**
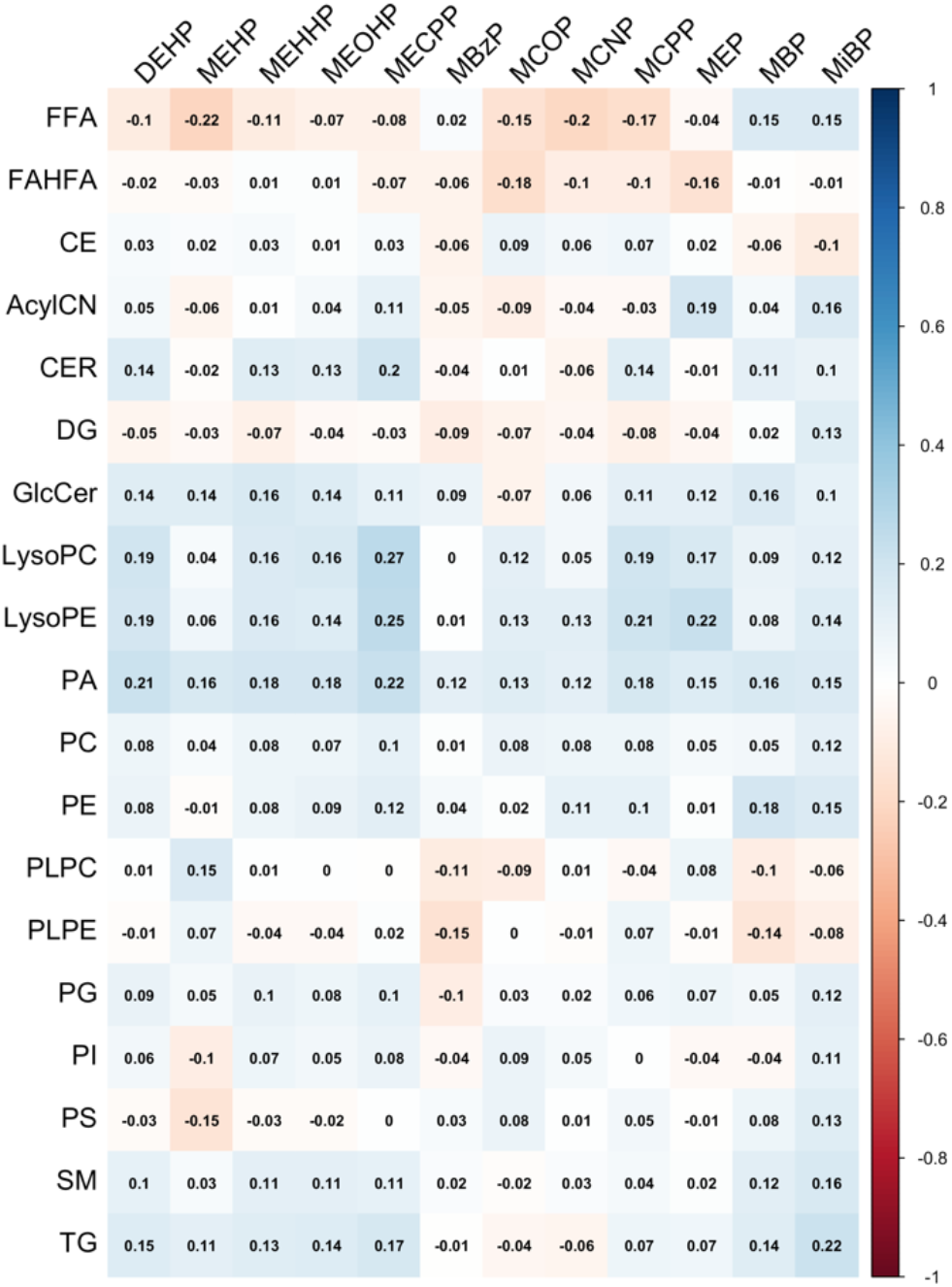
Correlation matrix between specific gravity corrected concentrations of phthalate metabolites and all lipid classes

**Figure 4.**
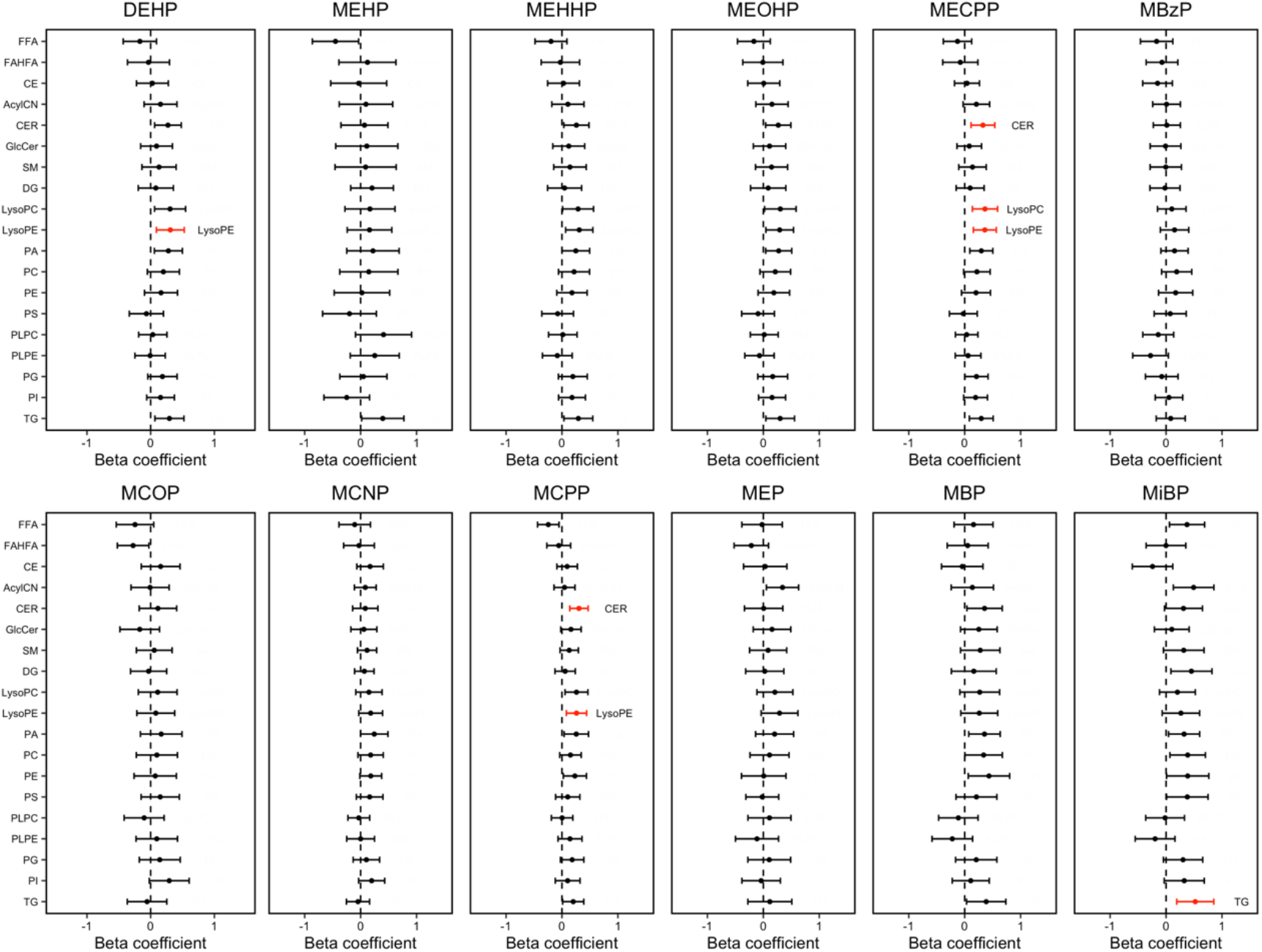
Percent change in lipid class sum z-score associated with urinary phthalate metabolite concentrations. Effect estimates are presented as percent change (%) for IQR increase in exposure biomarker concentration^ab^. Models were adjusted for maternal age, maternal education, fetal sex, pre-pregnancy BMI, and weight gain during pregnancy.

## DISCUSSION

Using a comprehensive lipidomics platform including almost six hundred lipid species, we explored the associations between phthalates and three hierarchical levels of lipid organization: (1) individual lipidome-wide metabolites, (2) subgroup clustering based on hydrocarbon chain saturation, and (3) whole lipid classes. We found evidence of associations between phthalates and each level of these organizational hierarchies of lipids. At the lipidome-wide level of individual metabolites, biomarkers of exposure to DEHP, MCPP, and MBP were associated with lipid metabolites from the DG family (DG 40:7) and lipids from the CER class. For lipid saturation subgroups, significant positive associations were observed for DEHP exposure biomarkers and saturated CER, mono-unsaturated CER, saturated LysoPC, mono-unsaturated LysoPC, and saturated TG, whereas MECPP was inversely associated with saturated FFA. When we evaluated the broadest level of whole lipid classes, a variety of phthalates are positively associated with CER, LysoPC, LysoPE, and TG lipid classes. The direction of associations indicated that higher concentrations of phthalates were largely associated with increased levels of all of the most strongly associated lipid classes. There was not substantive evidence of associations for phthalates in relation to whole lipid classes such as PA and PE, even though the analysis on the saturation subgroup level revealed associations with PA and PE lipids containing saturated and mono-unsaturated fatty acids.

We observed some variation in signatures from lipidome-wide analysis to lipid class analysis, and among distinct DG and PLPC classes. This supports the need to measure the associations between phthalates and lipids at an omics-scale to identify important specific lipid-derived biomarkers of maternal phthalate exposure on lipid disruption that would not be identified with less precise analyses of whole aggregate lipid classes. We generated a list of 20 individual lipids across several classes (DG, CER, LysoPE, PLPC, and FFA) that had robust significant associations after false discovery rate correction and may be used to uniquely characterize the lipidomic responses induced by exposure to different phthalate parent compounds (**Supplementary Table 1**). Among these lipids, DG 40:7 and PLPC 35:1 were the most significantly associated with exposure to multiple phthalates and may have high predictability for phthalate-induced disruption during pregnancy.

While there is some variation in signatures from lipidome-wide analysis to lipid class analysis, changes to the individual lipidome (represented by PLPC, LysoPE, CER lipids) related to phthalates (DEHP, DBP, and DiBP metabolites) were also observed in the changes of lipid-class and sub-class level analysis. For lipid classes including CER, LysoPC, LysoPE, and TG, we observed positive associations with ΣDEHP (LysoPE), MECPP (CER, LysoPC, LysoPE), MCPP (CER, LysoPE), and MiBP. These increases in lipid classes associated with DEHP metabolite concentrations are consistent with previous reports from animal studies showing that exposure to phthalates, particularly DEHP, influence circulating lipid levels and lipid signaling [17-20, 41]. In contrast, a recent study reported that embryonic exposure to DEHP and DBP in zebrafish significantly reduced the levels of individual lipids from the TG and DG classes [42].

Complementary enrichment analysis using proteomic data revealed biological processes associated with DBP (xenobiotic stimulus, lipoprotein metabolism, lipid catabolism, and regulation of FA and TG catabolism) and DEHP (lipase activity regulation, lipid absorption, lipid catabolism and metabolism) [42]. As noted in the introduction, one suggested mechanism is that phthalates, known as PPAR-agonists, are interfering with peroxisome-proliferator active receptor gamma (PPARγ) and therefore disrupting lipid metabolism [21, 22]. PPARγ is a member of the nuclear receptor superfamily that plays critical physiological roles regulating lipid metabolism [43, 44]. It is possible that PPARγ may serve as a mediator for maternal phthalate exposure and disruption of lipids. Previous studies have reported that PPARγ activation can increase both esterification as well as lipolysis [45, 46], which provided evidence for the increased levels of single chain lipid signals in our analysis, including the FFA, DG, and LysoPC, LysoPE families.

For lipid saturation subgroups, significant phthalate-induced lipid increases were exclusively observed for several lipid subgroups with saturated and mono-saturated carbon chains and DEHP metabolites (MEHHP, MEOHP, MECPP), MCPP, DBP metabolite (MBP), and DiBP metabolite (MiBP). The significance of the association was particularly related to the saturation of the lipid classes. All of the lipid groups in the above-mentioned associations contain saturated or mono-unsaturated hydrocarbon chains except for the associations between MCPP and poly-unsaturated CER. s Evidence from limited human studies have shown that increased levels of saturated and/or mono-unsaturated lipids can have negative effects on pregnancy and poor birth outcomes [47-49]. Therefore, the positive signals we observed between phthalates and saturated and mono-unsaturated lipid classes may be a proxy mechanism or mediator for suspected adverse effects of phthalates on pregnancy outcomes. On the other hand, the beneficial effects of the poly-unsaturated fatty acids were widely studied in previous literature [49, 50]. Formal mediation studies will contribute towards better understanding the potential for poly-unsaturated fatty acids to diminish the toxic effects of phthalates.

Previous research on this population has found that MBP and MiBP, biomarkers for DBP and DiBP exposure, were associated with increased odds of preterm birth, and urinary metabolites measured later in the pregnancy were more strongly associated with preterm birth [7]. Two other studies in Boston [8] and Mexico City [51] have also reported an association between DBP and DiBP metabolites and spontaneous preterm birth in pregnant women. Changes in the lipid classes such as FFA, CE, DG, and PLPE, have been related to alteration of gestational length in recently published work on the same Puerto Rico cohort [30]. Further, levels of multiple individual lipids and lipid classes (FFA, CER, PA, PE, and TG) were associated with phthalate metabolites MBP and MiBP in the present work. While our findings need to be confirmed, these findings support the hypothesis that disruption of lipid levels may be a possible pathway mediating phthalates and adverse pregnancy outcomes.

Our study employed a lipidome-wide analysis in relation to maternal phthalate exposure, while previous studies of the relationship between phthalate concentrations and lipids have been limited to animal studies or studies on targeted lipids such as triglyceride, LDL, and HDL. The extensive repertoire of lipid metabolites that were measured within the maternal lipidome is an important extension of previous research, as this provides an unprecedented opportunity to consider the maternal plasma lipidome in the context of maternal-neonate health. Importantly, this study contributes to the evolving evidence on the disrupting role of phthalates, most notably with DEHP metabolites on lipids, and is among the first to present these associations on a lipidomic scale.

There are a few limitations of this study. The analysis was limited by a fairly small sample size. While the results of this exploratory analysis are biologically plausible and supported by evidence derived from animal studies showing similar results, confirmation in larger diverse cohorts will be required in the future. Because of the cross-sectional nature of the study, windows of vulnerability during pregnancy were not evaluated. Given that phthalate exposure and lipid profiles change across the duration of pregnancy, it is important for future longitudinal studies to evaluate the temporality of the associations between phthalates and maternal lipidome throughout gestation. Although our analyses were adjusted for important potential confounders, residual confounding due to unmeasured variables (e.g., dietary factors) may still exist that could alternatively explain, at least in part, the observed associations.

## Conclusions

We employed a semi-targeted lipidomics analysis with more than five hundred measured metabolites in maternal plasma samples, which provided an opportunity to examine the relationship between urine phthalate metabolite concentrations with the plasma lipidome. We reported associations of maternal exposure to DEHP, DOP, DBP, and DiBP on omics scale plasma lipids and lipid classes among pregnant women, particularly with those containing saturated and monounsaturated fatty acid chains. We identified new potential lipidomic markers of phthalate-induced disruption during pregnancy. These results provide new insights into the relationship between maternal phthalate exposure and lipidomic profiles during gestation.

## Supporting information

Supplemental Results

## Data Availability

Data utilized for this analysis can be obtained by reasonable request by contacting the corresponding author (JDM)

## Data availability

### Disclaimer

The findings and conclusions in this report are those of the authors and do not necessarily represent the official position of the NIEHS, EPA, or Centers for Disease Control and Prevention (CDC). Use of trade names is for identification only and does not imply endorsement by the CDC, the Public Health Service, or the US Department of Health and Human Services.

## Author Contributions

P.A.: Statistical analysis; Investigation; Methodology; Writing, review and editing. M.T.A.: Writing, review and editing. D.J.W.: Conceptualization; Funding acquisition. B.M.: Conceptualization; Supervision; Funding acquisition. Z.R.: Data curation; Project administration. C.M.V.: Data curation; Project administration. A.A.: Conceptualization; Funding acquisition. J.F.C.: Conceptualization; Funding acquisition. J.D.M.: Conceptualization; Funding acquisition; Supervision.

## Funding

This study was supported by the Superfund Research Program of the National Institute of Environmental Health Sciences, National Institutes of Health (grants P42ES017198 and). Additional support was provided from NIEHS grant numbers P50ES026049, R01ES032203, and P30ES017885 and the Environmental influences on Child Health Outcomes (ECHO) program grant number UH3OD023251. Support for Max Aung was provided in part by NIH award P30ES030284.

## Acknowledgements

We thank the nurses and research staff who participated in cohort recruitment and follow up, as well as the Federally Qualified Health Centers (FQHC) and clinics in Puerto Rico who facilitated participant recruitment, including Morovis Community Health Center (FQHC), Prymed: Ciales Community Health Center (FQHC), Camuy Health Services, Inc. (FQHC), and the Delta OBGyn (Prenatal Clinic).

## Competing Interests

The authors declare that they have no financial or other conflict of interests.

## Notes

### Competing Interest Statement

The authors have declared no competing interest.

### Author Declarations

The research protocol was approved by the Ethics and Research Committees of the University of Puerto Rico and participating clinics, the University of Michigan, Northeastern University, and the University of Georgia.

