## Supplemental Results for "Maternal Urinary Phthalate Metabolites are Associated with Lipidomic Signatures Among Pregnant Women in Puerto Rico"

**This supplementary file contains 5 supplementary tables and 1 supplementary figure.**

**Supplementary Table 1.** Demographic Characteristics of n = 99 Pregnant Women from Puerto Rico.

| Variable | Mean (SD) | |
|---|---|---|
| Maternal Age | 26.5 (5.7) | |
| **Characteristic** | **Category** | **Count (Percent)** |
| Insurance Type | Private | 43 (43.4%) |
| | Public (mi salud) | 32 (32.3%) |
| | Missing | 24 (24.2%) |
| Maternal Education | <=High school/GED | 23 (23.2%) |
| | Some college or technical school | 31 (31.3%) |
| | College degree | 33 (33.3%) |
| | Master's degree or higher | 12 (12.1%) |
| | Missing | 0 (0%) |
| Household Income | <$10,000 | 30 (30.3%) |
| | ≥$10,000 to <$30,000 | 24 (24.2%) |
| | ≥$30,000 to <$50,000 | 20 (20.2%) |
| | ≥$50,000 | 10 (10.1%) |
| | Missing | 15 (15.2%) |
| Marital Status | Single | 22 (22.2%) |
| | Married or living together | 77 (77.8%) |
| | Missing | 0 (0%) |
| Gravidity (# Pregnancies) | 0 | 47 (47.5%) |
| | 1 | 33 (33.3%) |
| | >1 | 19 (19.2%) |
| | Missing | 0 (0%) |
| Pre-pregnancy BMI (kg/m$^2$) | ≤25 | 60 (60.6%) |
| | >25 to ≤30 | 19 (19.2%) |
| | >30 | 18 (18.2%) |
| | Missing | 2 (2%) |
| Employment Status | Unemployed | 55 (55.6%) |
| | Employed | 43 (43.4%) |
| | Missing | 1 (1%) |
| Smoking | Never | 86 (86.9%) |
| | Ever | 11 (11.1%) |
| | Current | 2 (2%) |
| | Missing | 0 (0%) |
| Exposure to Second-hand Smoking | None | 79 (79.8%) |
| | Up to 1 hour | 5 (5.1%) |
| | More than 1 hour | 11 (11.1%) |
| | Missing | 4 (4%) |
| Alcohol Consumption | None | 50 (50.5%) |
| | Before pregnancy | 41 (41.4%) |
| | Within the last few months | 6 (6.1%) |
| | Missing | 2 (2%) |

**Supplementary Table 2.** The top individual lipids associated with phthalate metabolites.

| Lipid names | Lipid class abbreviation | Phthalate | Parent phthalate | p value | q value |
|---|---|---|---|---|---|
| DG 40:7 | DG | MEHHP | DEHP | 4.07E-07 | 1.76E-04 |
| DG 40:7 | DG | MEOHP | DEHP | 1.48E-06 | 3.19E-04 |
| DG 40:7 | DG | ΣDEHP | DEHP | 4.25E-06 | 6.13E-04 |
| DG 40:7 | DG | MBP | DBP | 2.43E-05 | 2.62E-03 |
| DG 40:7 | DG | MEHP | DEHP | 3.43E-05 | 2.97E-03 |
| DG 40:7 | DG | MCPP | DOP | 5.46E-05 | 3.93E-03 |
| Cer NS 44:1 | CER | MCPP | DOP | 1.31E-05 | 5.99E-03 |
| LysoPE 18:0 | LysoPE | MECPP | DEHP | 1.28E-04 | 7.49E-03 |
| LysoPE 18:0 | LysoPE | MCPP | DOP | 1.39E-04 | 7.49E-03 |
| Cer NDS 36:0 | CER | MCPP | DOP | 5.00E-05 | 1.03E-02 |
| Cer NS 34:1 | CER | MCNP | DiDP | 7.09E-05 | 1.03E-02 |
| Cer NS 41:1 | CER | MCPP | DOP | 9.02E-05 | 1.03E-02 |
| DG 34:0 | DG | MECPP | DEHP | 1.94E-04 | 1.20E-02 |
| Cer NDS 40:0 | CER | MBP | DBP | 2.03E-04 | 1.38E-02 |
| Cer NDS 42:1 | CER | MCPP | DEHP | 2.57E-04 | 1.38E-02 |
| Cer NS 42:1 | CER | MCPP | DEHP | 2.57E-04 | 1.38E-02 |
| Cer NS 43:1 | CER | MCPP | DEHP | 2.60E-04 | 1.38E-02 |
| Cer NDS 42:0 | CER | MBP | DBP | 2.72E-04 | 1.38E-02 |
| PLPC 35:1 | PLPC | MiBP | DiBP | 2.11E-05 | 1.39E-02 |
| DG 36:1 | DG | MECPP | DEHP | 3.01E-04 | 1.62E-02 |
| DG 32:0 | DG | MECPP | DEHP | 3.93E-04 | 1.84E-02 |
| DG 34:0 | DG | ΣDEHP | DEHP | 4.69E-04 | 1.84E-02 |
| DG 34:0 | DG | MEOHP | DEHP | 5.09E-04 | 1.84E-02 |
| DG 40:7 | DG | MECPP | DEHP | 5.11E-04 | 1.84E-02 |
| Cer NDS 40:0 | CER | MiBP | DiBP | 5.28E-04 | 2.30E-02 |
| Cer NS 36:2 | CER | MiBP | DiBP | 5.55E-04 | 2.30E-02 |
| Cer NS 43:2 | CER | MCPP | DOP | 6.24E-04 | 2.34E-02 |
| Cer NS 41:3 | CER | MCPP | DOP | 6.66E-04 | 2.34E-02 |
| PLPC 35:1 | PLPC | MBP | DBP | 8.15E-05 | 2.69E-02 |
| PLPC 37:1 | PLPC | MECPP | DEHP | 1.46E-04 | 3.20E-02 |
| PLPC 35:1 | PLPC | MEHHP | DEHP | 2.57E-04 | 4.25E-02 |
| PLPC 35:1 | PLPC | MEOHP | DEHP | 3.59E-04 | 4.73E-02 |
| FFA 24:1 | FFA | MiBP | DiBP | 2.60E-04 | 5.00E-02 |

**Supplementary Table 3.** Lipid group classifications: lipidomics dataset was collapsed into groups of lipids based on the lipid class and the number of double bonds within the fatty acid tails. A list of the individual lipids within each group are reported.

| Lipid Group | Chain number | Number | Lipid Species |
|---|---|---|---|
| AcyICN | 1 | 7 | Acylcarnitine 14:1, Acylcarnitine 16:0, Acylcarnitine 18:0, Acylcarnitine 18:1, Acylcarnitine 18:2, Acylcarnitine 24:0 , Acylcarnitine 26:0 |
| CE-sat-mono | 1 | 5 | 16:0 CE, 18:0 CE, 16:1 CE, 17:1 CE, 18:1 CE |
| CE-poly | 1 | 9 | 18:2 CE, 18:3 CE, 20:2 CE, 20:3 CE, 20:4 CE, 22:4 CE, 20:5 CE, 22:5 CE, 22:6 CE |
| CER-sat | 1 | 10 | Cer[NDS] 34:0, Cer[NDS] 36:0, Cer[NDS] 38:0, Cer[NDS] 40:0, Cer[NDS] 41:0, Cer[NDS] 42:0, Cer[EODS] 53:0, Cer[NP] 34:0, Cer[NP] 40:0, Cer[NP] 42:0, |
| CER-mono | 1 | 16 | Cer[NS] 32:1, Cer[NS] 33:1, Cer[NS] 34:1, Cer[NS] 35:1, Cer[NS] 36:1, Cer[NS] 37:1, Cer[NS] 38:1, Cer[NS] 39:1, Cer[NS] 40:1, Cer[NS] 41:1, Cer[NS] 42:1, Cer[NS] 43:1, Cer[NS] 44:1, Cer[AS] 41:1, Cer[EODS] 49:1, Cer[NP] 42:1 |
| CER-poly | 1 | 12 | Cer[NS] 34:2, Cer[NS] 36:2, Cer[NS] 38:2, Cer[NS] 40:2, Cer[NS] 41:2, Cer[NS] 42:2, Cer[NS] 43:2, Cer[AS] 42:2, Cer[NDS] 42:2 , Cer[NS] 41:3, Cer[NS] 42:3, Cer[NS] 42:4 |
| DG-sat | 2 | 13 | DG 30:0, DG 31:0, DG 32:0, DG 33:0, DG 34:0, DG 36:0, DG 38:0, DG 30:1, DG 32:1, DG 33:1, DG 35:1, DG 36:1, DG 38:1 |
| DG-mono | 2 | 13 | DG 32:2, DG 33:2, DG 34:2, DG 35:2, DG 36:2, DG:37:2, DG 38:2, DG 34:3, DG 35:3, DG 36:3, DG 37:3, DG 38:3, DG 40:3 |
| DG-poly | 2 | 10 | DG 36:4, DG 38:4, DG 36:5, DG 38:5, DG 40:5, DG 38:6, DG 40:6, DG 38:7, DG 40:7, DG 40:8 |
| FFA-sat | 1 | 5 | FFA16:0, FFA18:0, FFA20:0, FFA22:0, FFA24:0 |
| FFA-mono | 1 | 4 | FFA18:1, FFA20:1, FFA22:1, FFA24:1 |
| FFA-poly | 1 | 7 | FFA18:2, FFA20:2, FFA22:2, FFA24:2, FFA22:3, FFA24:3, FFA20:4 |
| FAHFA | 1 | 2 | FAHFA 36:0, FAHFA 40:3 |
| GlcCer | 1 | 10 | GlcCer[NS] 34:1, GlcCer[NS] 36:1, GlcCer[NS] 38:1, GlcCer[NS] 40:1, GlcCer[NS] 41:1, GlcCer[NS] 42:1, GlcCer[NS] 43:1, GlcCer[NS] 34:2, GlcCer[NS] 41:2, GlcCer[NS] 42:2 |
| LysoPC-sat | 1 | 8 | LysoPC 14:0, LysoPC 15:0, LysoPC 16:0, LysoPC 18:0, LysoPC 19:0, LysoPC 20:0, LysoPC 22:0, LysoPC 24:0 |
| LysoPC-mono | 1 | 6 | LysoPC 16:1, LysoPC 17:1, LysoPC 18:1, LysoPC 19:1, LysoPC 20:1, LysoPC 24:1 |
| LysoPC-poly | 1 | 9 | LysoPC 18:2, LysoPC 20:2, LysoPC 18:3, LysoPC 20:3, LysoPC 20:4, LysoPC 22:4, LysoPC 20:5, LysoPC 22:5, LysoPC 22:6 |
| LysoPE-sat-mono | 1 | 4 | LysoPE 16:0, LysoPE 18:0, LysoPE 24:0, LysoPE 18:1 |

| | | | |
|---|---|---|---|
| LysoPE-poly | 1 | 5 | LysoPE 18:2, LysoPE 20:3, LysoPE 20:4, LysoPE 22:5, LysoPE 22:6 |
| PA-sat-mono | 2 | 4 | PA 32:1, PA 34:1, PA 34:2, PA 36:0 |
| PA-poly | 2 | 2 | PA 36:4, PA 38:6 |
| PC-sat | 2 | 23 | PC 24:0, PC 26:0, PC 29:0, PC 30:0, PC 31:0, PC 32:0, PC 33:0, PC 35:0, PC 36:0, PC 40:0, PC 28:1, PC 30:1, PC 31:1, PC 32:1, PC 33:1, PC 34:1, PC 35:1, PC 36:1, PC 37:1, PC 38:1, PC 39:1, PC 40:1, PC 42:1 |
| PC-mono | 2 | 22 | PC 30:2, PC 31:2, PC 32:2, PC 33:2, PC 34:2, PC 35:2, PC 36:2, PC 37:2, PC 38:2, PC 40:2, PC 42:2, PC 44:2, PC 30:3, PC 32:3, PC 33:3, PC 34:3, PC 35:3, PC 36:3, PC 37:3, PC 38:3, PC 39:3, PC 40:3, |
| PC-poly | 2 | 42 | PC 40:10, PC 42:10, PC 32:4, PC 34:4, PC 35:4, PC 36:4, PC 37:4, PC 38:4, PC 39:4, PC 40:4, PC 41:4, PC 42:4, PC 44:4, PC 34:5, PC 35:5, PC 36:5, PC 37:5, PC 38:5, PC 39:5, PC 40:5, PC 41:5, PC 42:5, PC 44:5, PC 35:6, PC 36:6, PC 37:6, PC 38:6, PC 39:6, PC 40:6, PC 41:6, PC 42:6, PC 35:7, PC 36:7, PC 37:7, PC 38:7, PC 39:7, PC 40:7, PC 41:7, PC 42:7, PC 39:8, PC 40:8, PC 42:8,PC 42:9 |
| PE-sat | 2 | 12 | PE 30:0, PE 32:0, PE 33:0, PE 34:0, PE 35:0, PE 36:0, PE 32:1, PE 33:1, PE 34:1, PE 35:1, PE 36:1, PE 38:1 |
| PE-mono | 2 | 12 | PE 32:2, PE 33:2, PE 34:2, PE 35:2, PE 36:2, PE 37:2, PE 38:2, PE 34:3, PE 35:3, PE 36:3, PE 37:3, PE 38:3 |
| PE-poly | 2 | 21 | PE 42:10, PE 34:4, PE 35:4, PE 36:4, PE 37:4, PE 38:4, PE 39:4, PE 40:4, PE 36:5, PE 37:5, PE 38:5, PE 40:5, PE 36:6, PE 37:6, PE 38:6, PE 39:6, PE 40:6, PE 38:7, PE 40:7, PE 40:8, PE 42:8 |
| PG-sat | 2 | 4 | PG 33:0, PG 36:0, PG 34:1, PG 36:1 |
| PG-mono | 2 | 3 | PG 34:2, PG 36:2, PG 36:3 |
| PI-sat | 2 | 3 | PI 32:0, PI 34:1, PI 36:1 |
| PI-mono | 2 | 4 | PI 34:2, PI 36:2, PI 36:3, PI 38:3 |
| PI-poly | 2 | 6 | PI 36:4, PI 38:4, PI 38:5, PI 40:5, PI 38:6, PI 40:6 |
| PS | 2 | 2 | PS 36:0, PS 38:4 |
| PLPC-sat | 2 | 15 | PLPC 24:0, PLPC 30:0, PLPC 32:0, PLPC 34:0, PLPC 37:0, PLPC 40:0, PLPC 42:0, PLPC 32:1, PLPC 34:1, PLPC 35:1, PLPC 36:1, PLPC 37:1, PLPC 38:1, PLPC 40:1, PLPC 42:1 |
| PLPC-mono | 2 | 17 | PLPC 32:2, PLPC 34:2, PLPC 35:2, PLPC 36:2, PLPC 37:2, PLPC 40:2, PLPC 42:2, PLPC 34:3, PLPC 35:3, PLPC 36:3, PLPC 37:3, PLPC 38:3, PLPC 39:3, PLPC 40:3, PLPC 41:3, PLPC 42:3, PLPC 44:3 |
| PLPC-poly | 2 | 23 | PLPC 36:4, PLPC 37:4, PLPC 38:4, PLPC 39:4, PLPC 40:4, PLPC 42:4, PLPC 44:4, PLPC 46:4, PLPC 36:5, PLPC 38:5, PLPC 39:5, PLPC 40:5, PLPC 41:5, PLPC 42:5, PLPC 44:5, |

| | | | |
|---|---|---|---|
| | | | PLPC 36:6, PLPC 37:6, PLPC 38:6, PLPC 39:6, PLPC 40:6, PLPC 41:6, PLPC 42:6, PLPC 44:6 |
| PLPE-sat | 2 | 10 | PLPE 32:0, PLPE 34:0, PLPE 37:0, PLPE 32:1, PLPE 33:1, PLPE 34:1, PLPE 35:1, PLPE 36:1, PLPE 38:1, PLPE 40:1 |
| PLPE-mono | 2 | 13 | PLPE 32:2, PLPE 33:2, PLPE 34:2, PLPE 35:2, PLPE 36:2, PLPE 37:2, PLPE 38:2, PLPE 40:2, PLPE 34:3, PLPE 36:3, PLPE 37:3, PLPE 38:3, PLPE 40:3 |
| PLPE-poly | 2 | 20 | PLPE 34:4, PLPE 35:4, PLPE 36:4, PLPE 37:4, PLPE 38:4, PLPE 39:4, PLPE 40:4, PLPE 42:4, PLPE 36:5, PLPE 38:5, PLPE 40:5, PLPE 42:5, PLPE 36:6, PLPE 37:6, PLPE 38:6, PLPE 39:6, PLPE 40:6, PLPE 41:6, PLPE 42:6, PLPE 44:6 |
| SM-sat | 2 | 23 | SM 30:0, SM 32:0, SM 34:0, SM 35:0, SM 36:0, SM 37:0, SM 38:0, SM 42:0, SM 30:1, SM 31:1, SM 32:1, SM 33:1, SM 34:1, SM 36:1, SM 37:1, SM 38:1, SM 39:1, SM 40:1, SM 41:1, SM 42:1, SM 43:1, SM 44:1, SM 45:1 |
| SM-mono | 2 | 25 | SM 30:2, SM 32:2, SM 33:2, SM 34:2, SM 35:2, SM 36:2, SM 37:2, SM 38:2, SM 39:2, SM 40:2, SM 41:2, SM 42:2, SM 43:2, SM 44:2, SM 34:3, SM 36:3, SM 37:3, SM 38:3, SM 39:3, SM 40:3, SM 41:3, SM 42:3, SM 43:3, SM 44:3, SM 45:3 |
| SM-poly | 2 | 20 | SM 36:4, SM 38:4, SM 40:4, SM 42:4, SM 43:4, SM 44:4, SM 47:4, SM 38:5, SM 40:5, SM 41:5, SM 42:5, SM 43:5, SM 44:5, SM 40:6, SM 41:6, SM 42:6, SM 44:6, SM 42:7, SM 44:7, SM 45:9 |
| TG-sat | 3 | 50 | TG 36:0, TG 38:0, TG 39:0, TG 40:0, TG 41:0, TG 42:0, TG 43:0, TG 44:0, TG 45:0, TG 46:0, TG 47:0, TG 48:0, TG 49:0, TG 50:0, TG 53:0, TG 54:0, TG 56:0, TG 40:1, TG 42:1, TG 43:1, TG 44:1, TG 45:1, TG 46:1, TG 47:1, TG 48:1, TG 49:1, TG 50:1, TG 51:1, TG 52:1, TG 53:1, TG 54:1, TG 55:1, TG 56:1, TG 58:1, TG 62:1, TG 42:2, TG 44:2, TG 46:2, TG 47:2, TG 48:2, TG 49:2, TG 50:2, TG 51:2, TG 52:2, TG 53:2, TG 54:2, TG 55:2, TG 56:2, TG 58:2 |
| TG-mono | 3 | 33 | TG 42:3, TG 44:3, TG 46:3, TG 48:3, TG 49:3, TG 50:3, TG 51:3, TG 52:3, TG 53:3, TG 54:3, TG 55:3, TG 56:3, TG 57:3, TG 58:3, TG 46:4, TG 48:4, TG 50:4, TG 51:4, TG 52:4, TG 53:4, TG 54:4, TG 55:4, TG 56:4, TG 58:4, TG 48:5, TG 50:5, TG 51:5, TG 52:5, TG 53:5, TG 54:5, TG 55:5, TG 56:5, TG 58:5 |
| TG-poly | 3 | 23 | TG 50:6, TG 50:6, TG 52:6, TG 54:6, TG 56:6, TG 54:7, TG 56:7, TG 58:7, TG 54:8, TG 55:8, TG 56:8, TG 58:8, TG 56:9, TG 58:9, TG 56:10, TG 58:10, TG 60:10, TG 58:11, TG 60:11, TG 60:12, TG 62:12, TG 62:14 |

**Supplementary Table 4.** Percent change in lipid subgroup sum z-score associated with urinary phthalate and metabolite concentrations. Effect estimates presented as percent change (%) for IQR increase in exposure biomarker concentration[ab]. Models were adjusted for maternal age, maternal education, fetal sex, pre-pregnancy BMI, weight gain during pregnancy.

| Lipid class | | | Phthalate | Change in lipid z-score | |
|---|---|---|---|---|---|
| **Abbreviation** | **Full name** | **Saturation** | | **IQR (95% CI)** | q value |
| **LysoPE** | lysophosphatidylethanolamine | saturated, monounsaturated | MECPP | 0.42 (0.22, 0.62) | 0.001 |
| **LysoPE** | lysophosphatidylethanolamine | saturated, monounsaturated | MCPP | 0.38 (0.19, 0.57) | 0.002 |
| **PE** | phosphatidylethanolamine | saturated | MBP | 0.69 (0.34, 1.03) | 0.003 |
| **LysoPE** | lysophosphatidylethanolamine | saturated, monounsaturated | DEHP | 0.38 (0.16, 0.59) | 0.004 |
| **CER** | ceramides | monounsaturated | MCPP | 0.31 (0.14, 0.48) | 0.01 |
| **LysoPE** | lysophosphatidylethanolamine | saturated, monounsaturated | MEHHP | 0.38 (0.15, 0.61) | 0.01 |
| **PC** | phosphatidylcholine | saturated | MiBP | 0.75 (0.33, 1.17) | 0.01 |
| **CER** | ceramides | saturated | MBP | 0.51 (0.22, 0.80) | 0.02 |
| **LysoPE** | lysophosphatidylethanolamine | saturated, monounsaturated | MEOHP | 0.36 (0.12, 0.6) | 0.02 |
| **CER** | ceramides | monounsaturated | MECPP | 0.34 (0.13, 0.56) | 0.02 |
| **CER** | ceramides | monounsaturated | MBP | 0.42 (0.11, 0.73) | 0.03 |
| **CER** | ceramides | monounsaturated | MEHHP | 0.30 (0.09, 0.52) | 0.03 |
| **CER** | ceramides | monounsaturated | MEOHP | 0.31 (0.09, 0.53) | 0.03 |
| **CER** | ceramides | monounsaturated | DEHP | 0.30 (0.09, 0.51) | 0.03 |
| **CER** | ceramides | polyunsaturated | MCPP | 0.27 (0.11, 0.43) | 0.03 |
| **PE** | phosphatidylethanolamine | monounsaturated | MCPP | 0.31 (0.12, 0.49) | 0.03 |
| **PE** | phosphatidylethanolamine | monounsaturated | MBP | 0.55 (0.19, 0.91) | 0.03 |
| **PA** | phosphatidic acid | saturated, monounsaturated | MBP | 0.4 (0.14, 0.67) | 0.03 |
| **PA** | phosphatidic acid | saturated, monounsaturated | MECPP | 0.29 (0.09, 0.49) | 0.03 |
| **PA** | phosphatidic acid | saturated, monounsaturated | MEOHP | 0.31 (0.09, 0.54) | 0.03 |
| **PA** | phosphatidic acid | saturated, monounsaturated | DEHP | 0.3 (0.08, 0.51) | 0.03 |
| **FFA** | free fatty acid | monounsaturated | MiBP | 0.47 (0.18, 0.76) | 0.03 |
| **FFA** | free fatty acid | saturated | MCPP | -0.31 (-0.5, -0.12) | 0.03 |

| | | | | | |
|---|---|---|---|---|---|
| **CER** | ceramides | saturated | MCPP | 0.38 (0.13, 0.64) | 0.04 |
| **TG** | triacylglycerol | saturated | MBP | 0.53 (0.18, 0.88) | 0.04 |
| **TG** | triacylglycerol | saturated | MECPP | 0.29 (0.1, 0.48) | 0.04 |
| **PA** | phosphatidic acid | saturated, monounsaturated | MEHHP | 0.3 (0.07, 0.53) | 0.04 |
| **PA** | phosphatidic acid | saturated, monounsaturated | MiBP | 0.36 (0.07, 0.65) | 0.04 |
| **LysoPC** | lysophosphatidylcholine | saturated | MECPP | 0.36 (0.14, 0.58) | 0.04 |
| **TG** | triacylglycerol | saturated | MEHHP | 0.29 (0.06, 0.51) | 0.04 |
| **TG** | triacylglycerol | saturated | MEOHP | 0.29 (0.07, 0.51) | 0.04 |
| **TG** | triacylglycerol | saturated | MiBP | 0.53 (0.12, 0.93) | 0.04 |
| **TG** | triacylglycerol | saturated | DEHP | 0.28 (0.08, 0.48) | 0.04 |
| **LysoPC** | lysophosphatidylcholine | monounsaturated | MECPP | 0.38 (0.14, 0.62) | 0.04 |
| **CER** | ceramides | saturated | MECPP | 0.32 (0.08, 0.56) | 0.04 |
| **CER** | ceramides | saturated | MiBP | 0.48 (0.13, 0.83) | 0.04 |

**Supplementary Table 5.** Percent change in lipid group sum z-score associated with urinary phthalate and metabolite concentrations. Effect estimates presented as percent change (%) for IQR increase in exposure biomarker concentration[ab]. Models were adjusted for maternal age, maternal education, fetal sex, pre-pregnancy BMI, weight gain during pregnancy.

| **Lipid class** | **Lipid class full name** | **Phthalate** | **Change in lipid z-score IQR (95% CI)** | **q value** |
|---|---|---|---|---|
| CER | ceramides | MCPP | 0.31 (0.14,0.46) | 0.008 |
| LysoPE | lysophosphatidylethanolamine | MECPP | 0.36 (-.15, 0.56) | 0.02 |
| CER | ceramides | MECPP | 0.32 (0.11, 0.54) | 0.03 |
| LysoPC | lysophosphatidylcholine | MECPP | 0.36 (0.14, 0.59) | 0.04 |
| LysoPE | lysophosphatidylethanolamine | MCPP | 0.26 (0.08, 0.44) | 0.04 |
| LysoPE | lysophosphatidylethanolamine | DEHP | 0.31 (0.09, 0.53)] | 0.04 |
| TG | triacylglycerol | MiBP | 0.52 (0.19, 0.85) | 0.05 |

**Supplementary Figure 1**. Euler diagram showing the overlap between individual lipids associated with urinary phthalate metabolite concentrations, when categorized by phthalate parent compound. Models were adjusted for maternal age, maternal education, fetal sex, pre-pregnancy BMI, weight gain during pregnancy.

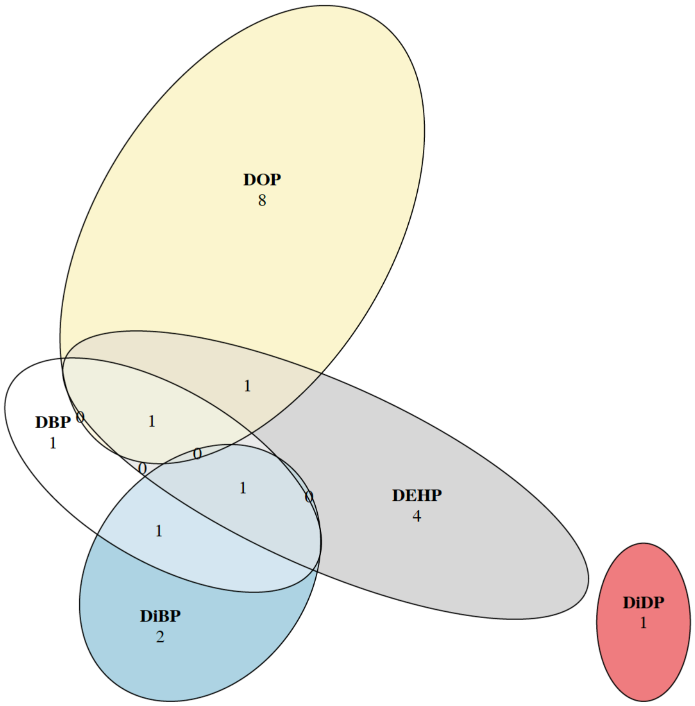
